# Constructing Tumor Immune Microenvironment and Identifying the Immune-Related Prognostic Signatures in Colorectal Cancer Using Multi-omics Data

**DOI:** 10.1101/2021.08.30.21262762

**Authors:** Shuai Zhang, Jiali Lv, Bingbing Fan, Zhe Fan, Chunxia Li, Bingbing Gu, Wenhao Yu, Tao Zhang

**Author notes:** **Correspondence & Reprints**: Tao Zhang, MD, PhD, Department of Biostatistics, School of Public Health, Cheeloo College of Medicine, Shandong University, Jinan, Shandong, 250012, China, PO Box 100, 44 Wenhuaxi Rd, Jinan 250012, China. These authors made equally contributions (Shuai Zhang, Jiali Lv).

## Abstract

**Background:** The tumor immune microenvironment (TIME) plays a key role in occurrence, progression and prognosis of colorectal cancer (CRC). However, the genetic and epigenetic alterations and potential mechanisms in the TIME of CRC are still unclear.

**Methods:** We investigated the immune-related differences in three types of genetic or epigenetic alterations (gene expression, somatic mutation, and DNA methylation) and considered the potential roles that these alterations have in the immune response and the immune-related biological processes by analyzing the multi-omics data from The Cancer Genome Atlas (TCGA) portal. Additionally, a four-step method based on LASSO regression and Cox regression was used to construct the prognostic prediction model. Cross validation was performed to validate the model.

**Results:** A total of 1,745 differentially expressed genes, 178 differentially mutated genes and 1,961 differentially methylation probes were identified between the high-immunity group and the low-immunity group. We retained 15 genetic and epigenetic variables after using LASSO regression and Cox regression. For the prognostic predictions on the TCGA profiles, the performance of the model with 1-year, 3-year, and 5-year areas under the curve (AUCs) equal to 0.861, 0.797, and 0.875. Finally, the overall risk score model was constructed based on genetic, epigenetic, demographic and clinical characteristics, which comprised 18 variables and achieved a high degree of accuracy on the 1-year (AUC = 0.865), 3-year (AUC = 0.839), and 5-year (AUC = 0.914) survival predictions. Kaplan-Meier survival analysis demonstrated that the overall survival of the high-risk group was significantly poorer compared with the low-risk group. Prognostic nomogram, calibration plot and cross validation showed excellent predictive performance.

**Conclusions:** Our study provides a new clue to explore the TIME of CRC patients in genetic and epigenetic aspects. Meanwhile, the prognostic model also has clinical prognostic value and may provide new indicators for the treatment of CRC patients.

## INTRODUCTION

Colorectal cancer (CRC) is currently the third most common cancer in terms of morbidity and mortality according to Cancer Statistics 2020. It results in approximately 147,950 new cases and 53,200 deaths in the United States each year^1^. The tumor-node-metastasis (TNM) staging system proposed by The international American Joint Committee on Cancer (AJCC) provides both traditional and current guidelines for the classification of CRC patients^2,3^. However, even patients in the same stage and receiving the same clinical treatment have different survival outcomes^4,5^. Therefore, it is very crucial to find new indicators with prognostic value for the treatment of patients.

With the development of medical technology, the treatment of CRC patients has developed from surgery, chemotherapy and radiotherapy to the current biological targeted therapy and immunotherapy^6,7^. For example, immune checkpoint blockers (ICBs) therapies show satisfactory results in the treatment of patients with microsatellite instability-high (MSI-H)/mismatch repair deficiency (dMMR)^8^. However, this method is affected by the tumor immune microenvironment (TIME). Therefore, an in-depth understanding of the tumor microenvironment (TME) is of great significance to the choice of immunotherapy.

TME refers to the surrounding microenvironment in which tumor cells exist, including blood vessels, immune cells, fibroblasts, bone marrow-derived inflammatory cells, various signal molecules and extracellular matrix, which are closely related to tumor progression and immunotherapy results^9-11^. Tumor-associated macrophages (TAMs) are major components of tumor microenvironment that frequently associated with tumor metastasis and chemoresistance^12,13^. In addition, some indicators related to the TIME are gradually established. For example, 5 genes were identified as fibroblast-specific biomarkers in TIME of poorer prognosis of CRC patients by Zhou et al.^14^. Additionally, Wang et al.^15^ established an immune-related prognostic signature consisting of 8 genes which reflects the dysregulated TIME and has a potential for better CRC patient management. However, most of the prognostic models for CRC only contain gene expression profiles and cannot reflect the overall situation of TIME^15,16^.

To solve this problem, we aim to use the gene expression profiles from The Cancer Genome Atlas (TCGA) portal to construct TME infiltration pattern and evaluate the relationship between TIME and the genetic or epigenetic signatures through different immune levels. We expected to construct a prognostic prediction model using significant gene expression, whole-exome sequencing and DNA methylation data in order to have a comprehensive understanding of the immune-related genetic and epigenetic changes in CRC patients.

## MATERIALS AND METHODS

### Data Collection

Totally, 428 tumor samples and 38 normal samples of RNA-seq data, 363 tumor profiles of somatic mutation data processed by VarScan2, 271 tumor DNA methylation profiles based on the Illumina Human Methylation 450 platform, and corresponding demographic and clinical information were collected from the TCGA portal.

### TME Construction

Construction of the TME was based on the transcriptome profiles with HTSeq-FPKM format from TCGA portal using the estimate algorithm^16^. Stromal, immune and estimate scores were calculated by R package “estimate” and compared between tumor samples and normal samples. Then, X-tile software^17^ was used to determine the best cut-off values of stromal, immune and estimate scores, the tumor samples were divided into high-immunity group and low-immunity group by the best cut-off value of immune scores. CIBERSORT algorithm^18^ was used to quantify the proportions of 22 various immune cell components of CRC patients based on RNA-seq data. The *P* value of CIBERSORT represented the statistical significance, and samples with a *P* value < 0.05 were subjected to subsequent correlation analysis. Mann-Whitney U test or Kruskal-Wallis test was used to compare stromal, immune and estimate scores between different groups. For the tumor samples of the somatic mutation and DNA methylation data, we also constructed a high-immunity group and a low-immunity group by mapping the sample IDs of the gene expression profiles.

### Multi-omics Data Analysis

For gene expression profiles, calculation of differential expressed genes (DEGs) was conducted using R package “edgeR”^19^ between the high-immunity group (n = 322) and the low-immunity group (n = 106). The criteria of DEG was log2 |Fold Change| > 1 and false discovery rate (FDR) < 0.05.

For somatic mutation data, we obtained profiles disposed by Varscan2^20^ from TCGA database and analyzed via the R package “maftools”, which provides a large amount of commonly used analysis and visualization modules in cancer genomic studies^21^. Somatic mutation profiles of the high-immunity group (n = 267) and the low-immunity group (n = 96) were used to detect the mutation types and SNVs. Identification of differential mutated genes (DMGs) was based on Fisher’s exact test with a *P* value < 0.05. The co-occurrence and mutually exclusivity analysis were carried out separately on the high-immunity cohort and the low-immunity cohort based on the CoMEt algorithm^22^.

DNA methylation profiles based on the Illumina Human Methylation 450 platform were obtained from TCGA portal to detect the relationship between DNA methylation pattern and different immune levels. We processed DNA methylation data by deleting DNA methylation sites in which more than 20% samples have missing beta values and filling in remaining missing beta values based on k-nearest neighbor method. Then, R package “ChAMP”^23^ was used to filtered low-quality probes by ChAMP’s filter function, normalized by champ.norm function and detected differential methylation probes (DMPs) between the high-immunity group (n = 191) and the low-immunity group (n = 80) by champ.DMP function. Furthermore, the correlation between the DNA methylations and gene expressions in different immune levels was explored via the Pearson correlation.

### Functional Enrichment Analysis

Gene Ontology (GO) enrichment analysis of the DEGs and DMP-associated genes and Gene Set Enrichment Analysis (GSEA) were performed based on the R package “clusterProfiler”^24^.

### Construction and Validation of Prognostic Model

Multi-omics data, overall survival time and status were used to establish prognostic model. We only considered samples which had gene expression values, somatic mutation status and DNA methylation probes to construct prognostic prediction model.

The workflow of exploring the prognostic model from various immunity-related genetic or epigenetic alterations in CRC patients consists of four steps: (1) the LASSO Cox regression was performed to drop the less contributive variables using the R package “glmnet”^25^. (2) multivariate Cox proportional hazard regression with stepwise method to fit the prognostic prediction model using selected variables based on the “survival” R package and stepAIC function in the R package “MASS”. (3) The predictive risk score of each patient was calculated based on prognostic model by predict function in “survival” R package. The patients were stratified into high-risk group and low-risk group by the median value of their risk scores. Then, Kaplan-Meier method and the log-rank test was used to compare the difference in overall survival between high and low risk groups. 1-year, 3-year, and 5-year receiver operating characteristic curve (ROC) were drew using the R package “timeROC”^26^. Nomogram and calibration plot were predicted by “rms” and “caret” R package, respectively. (4) The prognostic signatures were constructed again combined with three other demographic and clinical factors (age, sex and pathologic stage) and further included in the multivariate Cox proportional hazards regression model to evaluate the overall effect and assess the performance as step 3.

For the validation of prognostic model, 5-fold cross validation was performed to accomplish reliable predictive measurement based on “caret” R package^27^. In the 5-fold cross validation, the TCGA samples were randomly divided into five different sets, four sets were used for training, and the remaining set was used for validation.

## RESULTS

### Construction of the TME

RNA-seq profiles with complete clinical information were collected from 466 samples, including 428 tumor samples and 38 normal samples. As shown in **Figure 1A**, the stromal scores ranged from -2,165.02 to 1,679.72, the immune scores were between -892.30 to 2,378.73, and the estimate scores were distributed from -2,843.32 to 4,058.45. Stromal, immune and estimate scores in normal group were significantly higher than those in tumor group (*P* < 0.05).

**Figure1.**
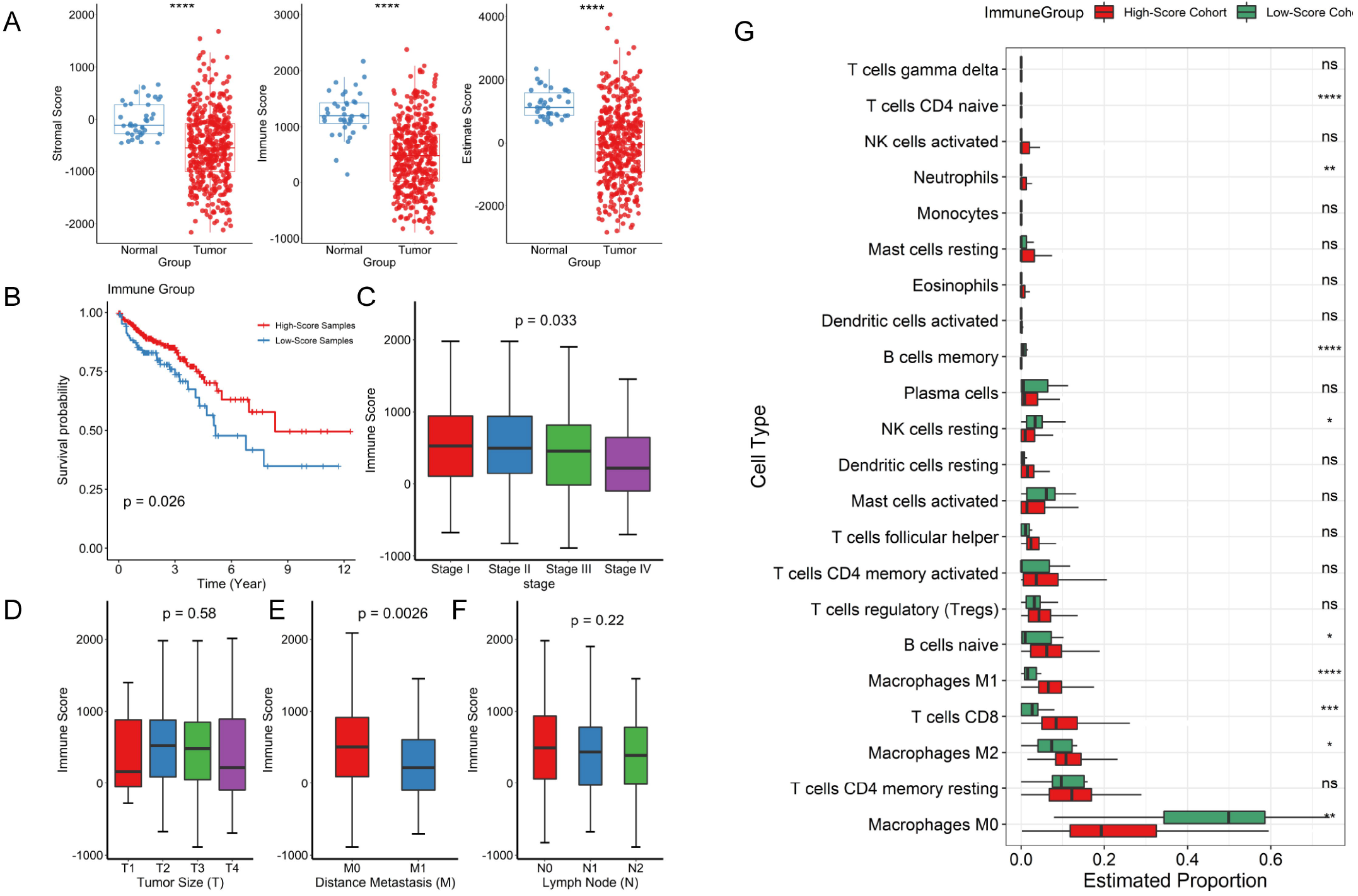
Construction of TME in CRC patients. (A) Comparison of the distributions of estimate, immune and stromal scores between tumor and normal samples. (B) Kaplan-Meier curves show the independent relevance between overall survival time and immune scores. (C) Comparison of immune scores on the pathological stage. Comparisons of immune scores on(D) tumor size, (E) distance metastasis, and (F) lymph nodes. (G) Comparisons of the immune cell members between high-immunity group and low-immunity group. Symbols indicated statistical significance for the Mann-Whitney U test: ns, *P* > 0.05; *, *P* <= 0.05; **, *P* <= 0.01;***, *P* <= 0.001; ****, *P* <= 0.0001.

Then, the tumor group was retained to further construct the TME. The best cut-off values of stromal, immune and estimate scores, which were -28.83, 14.66 and -1,451.13, respectively. The tumor group were divided into high and low score groups according to the best cut-off values. As shown in **Figure S1A**, there were no significant differences in stromal scores among pathologic stage (*P* = 0.77), tumor size (*P* = 0.30), lymph node (*P* = 0.13), and distance metastasis (*P* = 0.65). Kaplan–Meier plot revealed that survival probability between low stromal group and high stromal group had no significant difference (*P* = 0.095). The same situation was existed in estimate scores among pathologic stage (*P* = 0.59), tumor size (*P* = 0.59), lymph node (*P* = 0.67), distance metastasis (*P* = 0.08) and overall survival (*P* = 0.140) (**Figure S1B**). In contrast, the immune scores were different among pathologic stage (*P* < 0.05) (**Figure 1C**). Multiple comparisons showed that stage II had a higher immune score in comparison to stage IV (FDR < 0.05). M0 had a significantly higher immune score than M1 in distance metastasis (*P* < 0.05) (**Figure 1E**). Furthermore, samples with low immune scores (n = 106) had a bad prognosis compared with those subjects belong to high immune scores group (n = 322) (**Figure 1B**).

### Exploring the Tumor-infiltrating Immune Compositions

The immune infiltrating cell composition of each sample was shown in **Figure S2**, CD4 resting memory T cell, CD8 T cell, naive B cell and macrophages (including M0, M1, and M2) account for more than half proportions. Next, we compared the components of immune cells in the high and low-immunity groups, the results showed that the proportion of M1 macrophages, M2 macrophages, CD8 T cells, naive B cells, and neutrophils in the high immunity group was significantly larger, while the low immunity group had significantly larger fractions of M0 macrophages, memory B cells, resting NK cells, and naive CD4 T cells (*P* < 0.05) (**Figure 1G**).

### Identification of Differentially Expressed Genes Related to TIME

The collected RNA-seq profiles of CRC patients were used to identify differentially expressed genes (DEGs) between the high-immunity group and the low-immunity group. In the high-immunity group, 1617 genes and 128 genes were significantly up-regulated and down-regulated, respectively (**Figure 2A**).

**Figure2.**
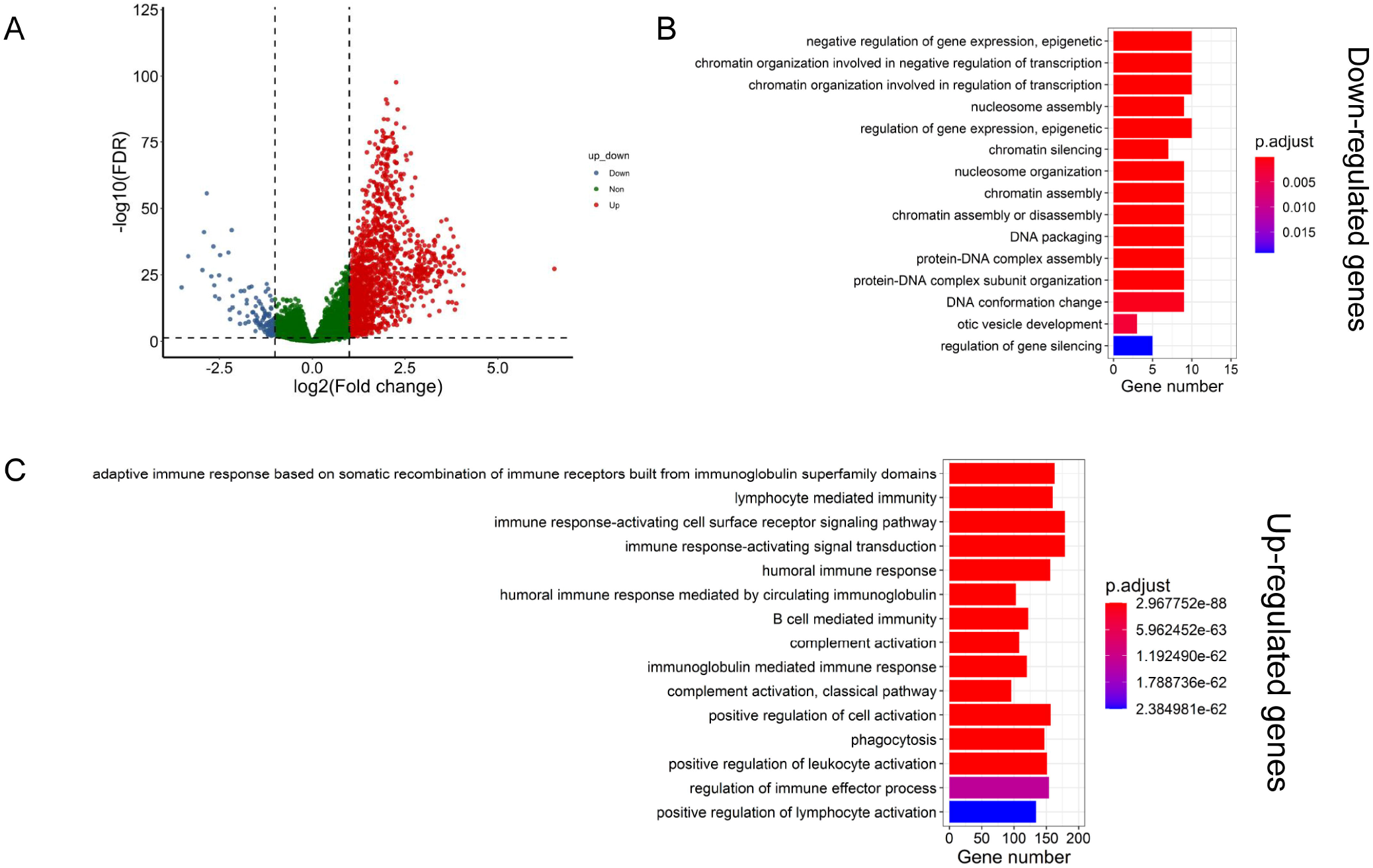
Investigation of the Immune Infiltration-Dependent Expression Change. (A) Volcano plot showing the differentially upregulated (red nodes) and downregulated genes (blue nodes). (B) Bar plot showing the biological processes enriched by the downregulated genes. (C) Bar plot showing the biological processes enriched by the upregulated genes. The y axis in (B) and (C) reflects the name of GO terms. The x axis reflects the overlapped gene numbers between each GO term and query gene set. The color of the bars represents the gradient of adjusted p values (FDR correction).

The results of Gene Ontology (GO) term enrichment analysis showed that the up-regulated genes were found to be enriched in the biological processes related to immune regulation and activation such as “lymphocyte mediated immunity” and “humoral immune response” (**Figure 2C**), meanwhile, the down-regulated genes were significantly enriched in the GO term such as “negative regulation of gene expression, epigenetic” and “nucleosome assembly”, indicating that those genes played a role in gene expression and transcription processes (**Figure 2B**).

### Landscape of Significantly Mutated Genes in Different Immune Levels

On the whole. As shown in **Figure 3A**, the numbers of nonsense mutation and nonstop mutation in high-immune group were larger than low-immune group (*P* < 0.05), and the percentage of nonstop mutation in high-immune cohort was larger than low-immune group (*P* < 0.05). Meanwhile, missense mutation was the largest proportion of gene mutations, accounting for almost 85% in both the high-immunity cohort and low-immunity group, and there appeared a larger percentage of missense mutations in the low-immunity group comparing to the high one (*P* < 0.05) (**Figure S3A**). The remaining mutation types showed no significant difference. As for SNVs, C > T mutation was the vast majority of various SNV types both in the high-immunity cohort (median frequency = 294.15; median percentage = 60.63%) and the low-immunity cohort (median frequency = 176.18; median percentage = 61.49%), the numbers of T > G, T > C and T > A, as well as the percentage of T > C in the high-immunity cohort were larger in comparison with the low-immunity cohort (**Figure 3B** and **Figure S3B**).

**Figure 3.**
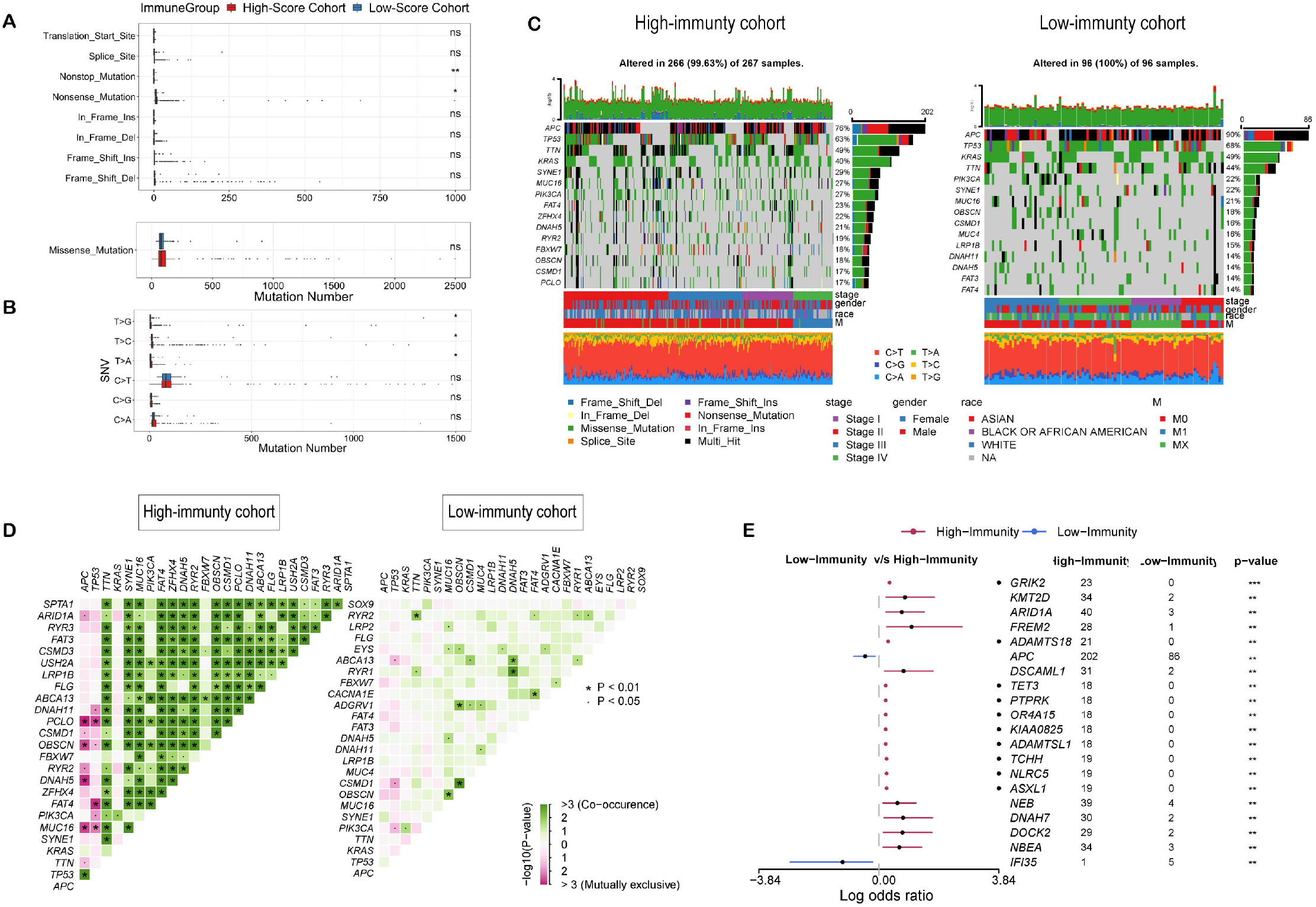
Landscape of Somatic Mutation in High-Immunity and Low-Immunity Cohorts. (A) Boxplots showing the comparisons of mutation frequencies of every mutation type classified by effects every mutation type classified by effects. (B) Boxplots showing the comparisons of mutation frequencies of every mutation type classified by effects every mutation type classified by effects and SNP. (C) Waterfall plot shows the mutation distribution of the top 15 most frequently mutated genes. The central panel shows the types of mutations in each CRC sample. The upper panel shows the mutation frequency of each CRC sample. The bar plots on the right side show the frequency and mutation type of genes mutated in the low-immunity and high-immunity cohort. The lower part shows the clinical features (pathological stage, gender, sex, and race) and SNV types of each sample. The bottom panel is the legend for mutation types and clinical features. (D) The heatmap illustrates the mutually co-occurring and exclusive mutations of the top 25 frequently mutated genes. The color and symbol in each cell represent the statistical significance of the exclusivity or co-occurrence for each pair of genes. (E) Forest plot displays the top 10 most significantly differentially mutated genes between two groups. Symbols indicated statistical significance for the Mann-Whitney U test or Fisher’s exact test: ns, *P* > 0.05; *, *P* <= 0.05; **, *P* <= 0.01; ***, *P* <= 0.001.

Panoramic views of the top 15 most frequently mutated genes of the high-immunity and the low-immunity groups in each sample were displayed in **Figure 3C**. APC, TP53, TTN and KRAS were accounted for the top four positions in both cohorts. As shown in **Figure 3D**, the number of pairwise genes of significant co-occurring and mutually exclusive mutations in the high immunity group is larger than that in the low immunity group. However, mutually exclusive relationship between TP53-PIK3CA was significantly in both two cohorts (*P* < 0.05). Additionally, there were 178 genes showed differential mutation frequencies between the high-immunity and the low-immunity cohorts (Fisher’s exact test, *P* < 0.05), ranked by ascending order of p-value (**Table S1**), and the top 20 genes are displayed in **Figure 3E**.

### Depicting Differential Methylation Probes in TIME

DNA methylation is one of the most ubiquitous epigenetic modifications regulating gene expression that have an impact on the progression of various cancers^28^. A total of 1,961 DMPs related to immune levels were identified based on the criteria of Δ β > 0.15 and FDR <0.05 (**Figure 4A and Table S2**). In the high-immune group, there were 155 (7.90%) hypermethylation sites related to 124 genes and were located on 133 CpG islands; 1806 (92.10%) hypomethylation sites involving 1370 genes and were located on 640 CpG islands. On the whole, the high-immunity group tends to have hypomethylated sites. Of the 1,473 DMP-associated genes, 21 genes had both hypermethylation sites and hypomethylation sites; 1349 genes had only hypomethylation sites, and 103 genes had only hypermethylation sites. Meanwhile, a lot of DMP-associated genes were differentially expressed between the high-immunity group and the low-immunity group according to the criteria of log2 |Fold Change|> 1 and FDR < 0.05. Compared to the low-immunity group, there were 144 up-regulated DEGs and a down-regulated DEG (GRM4) among 1370 hypomethylation genes (**Table S3**) and only 15 up-regulated DEGs among the 124 hypermethylation genes (**Table S4**) in the high-immunity group. The results of GO enrichment analysis showed that the DMP-associated genes were mainly enriched in the biological processes related to neuronal signal regulation such as “dendrite morphogenesis” and “synapse organization” (**Figure 4B**). Especially, gene set enrichment analysis (GSEA) shows that the DMP-associated genes with highly positive fold change differences have more essential contributions to the biological processes which related to immune regulation, indicating that DNA methylation may be involved in immune response through neuronal signal transmission (**Figure 4C**).

**Figure 4.**
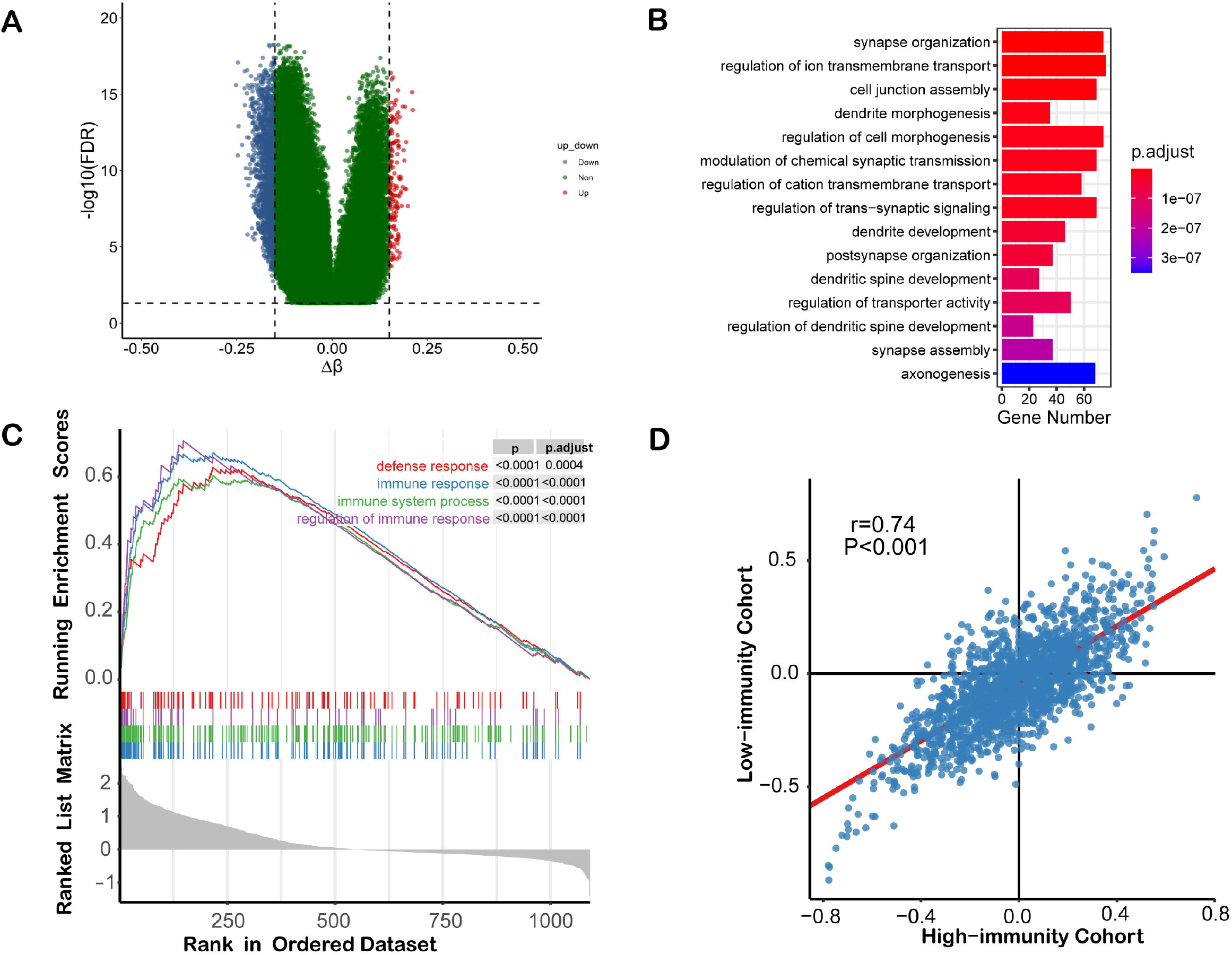
DNA Methylation Pattern in TIME. (A) Volcano plot showing the differentially methylation probes. (B) Bar plot showing the results of GO biological process enrichment analyses on DMP-associated genes. (C) Gene set enrichment analysis (GSEA) shows the significant enrichment in four immune-associated biological processes. (D) Scatterplot showing the correlations of methylation-expression correlation coefficients between high-immunity and low-immunity cohorts.

The results of correlation between DNA methylations and gene expressions in different immune levels showed that among the 1,473 identified DMP-associated genes, there were 111 positively correlated and 259 negatively correlated genes in the low-immunity group (**Table S5**). In contrast, in the high-immunity group, there were 336 positively correlated and 376 negatively correlated genes (**Table S6**) (Pearson correlation, *P* < 0.05). In general, the correlation between DNA methylations and gene expressions was more inclined to negatively correlated in the low-immunity group. Meanwhile, as shown in **Figure 4D**, the immune level did not affect the correlation between DNA methylations and gene expressions.

### Construction and Validation of Prognostic Model

In summary, there were some significant immune-related changes in the multi-omics data, including gene expressions, somatic mutations, and DNA methylations. In terms of gene expressions, a total of 1617 up-regulated genes and 128 down-regulated genes were detected in the high-immunity group. For somatic mutations, 10 and 168 significantly mutated genes were explored in the low-immunity group and the high-immunity group, respectively. As to DNA methylations, 1961 differential methylation probes related to immune levels were identified. We obtained 264 samples with these three types of data to construct prognostic model, detailed demographic and clinical information were displayed in **Table 1**.

**Table 1.**
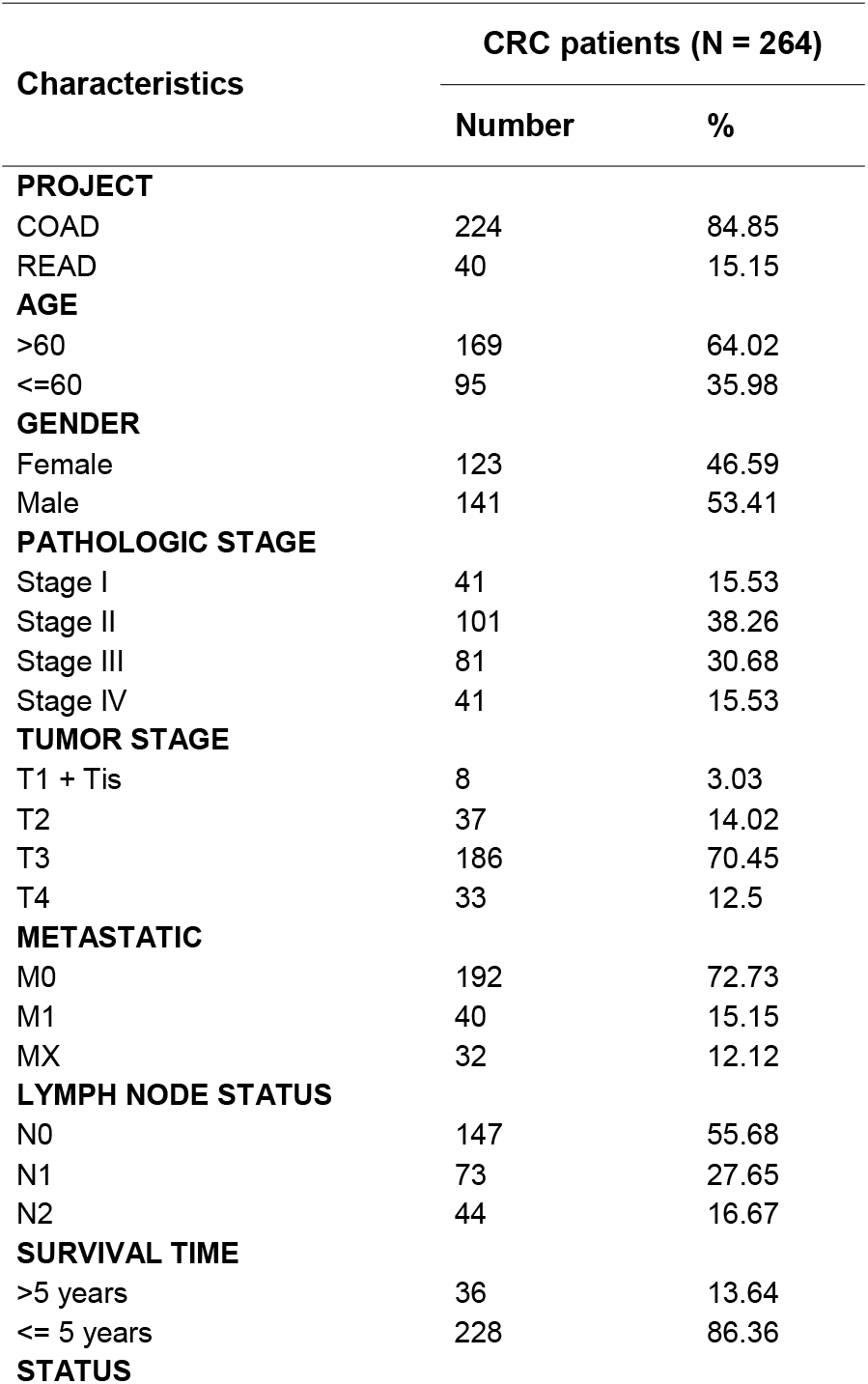
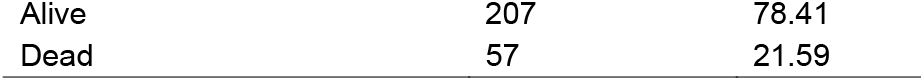
Summary information of colorectal cancer patients.

| Characteristics | CRC patients (N = 264) |  |
| --- | --- | --- |
|  | Number | % |
| **PROJECT** |  |  |
| COAD | 224 | 84.85 |
| READ | 40 | 15.15 |
| **AGE** |  |  |
| >60 | 169 | 64.02 |
| <=60 | 95 | 35.98 |
| **GENDER** |  |  |
| Female | 123 | 46.59 |
| Male | 141 | 53.41 |
| **PATHOLOGIC STAGE** |  |  |
| Stage I | 41 | 15.53 |
| Stage II | 101 | 38.26 |
| Stage III | 81 | 30.68 |
| Stage IV | 41 | 15.53 |
| **TUMOR STAGE** |  |  |
| T1 + Tis | 8 | 3.03 |
| T2 | 37 | 14.02 |
| T3 | 186 | 70.45 |
| T4 | 33 | 12.5 |
| **METASTATIC** |  |  |
| M0 | 192 | 72.73 |
| M1 | 40 | 15.15 |
| MX | 32 | 12.12 |
| **LYMPH NODE STATUS** |  |  |
| N0 | 147 | 55.68 |
| N1 | 73 | 27.65 |
| N2 | 44 | 16.67 |
| **SURVIVAL TIME** |  |  |
| >5 years | 36 | 13.64 |
| <= 5 years | 228 | 86.36 |
| **STATUS** |  |  |
| Alive | 207 | 78.41 |
| Dead | 57 | 21.59 |

We identified signatures related to prognosis from numerous genetic changes through LASSO regression and Cox proportional hazard regression. Firstly, the LASSO Cox regression was performed to drop the less contributive variables, the regularization parameter was set to log(lambda) = -2.94. 32 variables were reserved (**Table S7**). Then, 32 variables included in the stepwise multivariate Cox proportional hazard regression to fit the best model that comprising of 19 variables. We tested the collinearity between the variables and deleted 4 of them, leaving 15 variables as prognostic signatures, including the expression levels of AL139352.1, CST6, DNASE1L3, FCRL2, GRM4, HOTAIR, IGLV6-57, PLEKHF1 and SLC25A24P1, the mutation status of GRIK2 and NXF3, and the methylation levels of cg01726287, cg10717401, cg14332653 and cg16559243 (**Figure S5A**), the average concordance index (C-index) equal to 0.825 and the average AUC values predicted by the model for 1-year, 3-year, and 5-year were 0.861, 0.797, 0.875, respectively (**Figure 5A**). Next, we calculated the risk score of each sample according to the established model, the samples were divided into high-risk group and low-risk group according to the median risk score. Kaplan-Meier survival analysis (**Figure S5B**) displayed that the overall survival of the high-risk group was significantly poorer compared with the low-risk group (*P* < 0.001).

**Figure 5.**
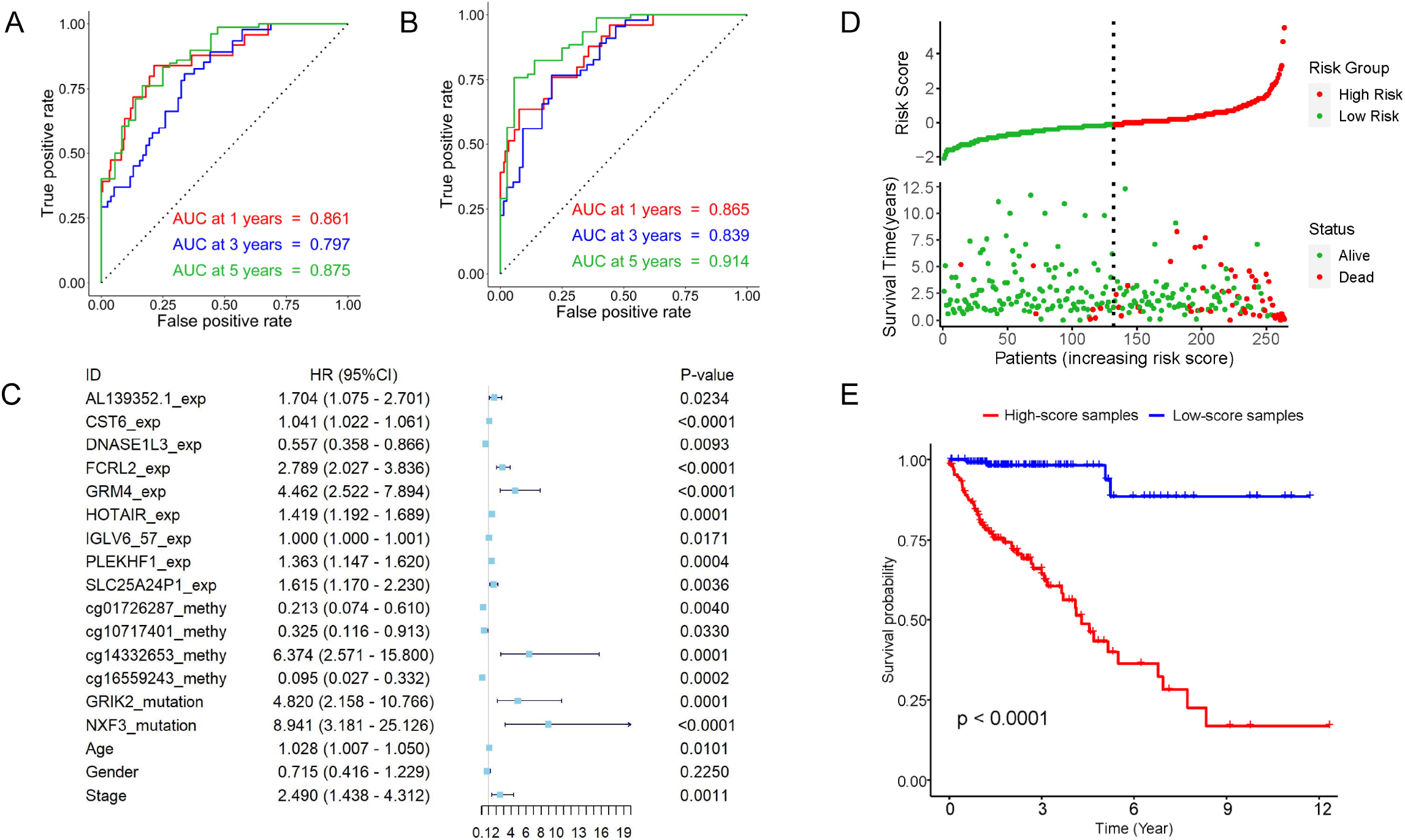
Establishing a Prognostic Model for CRC. (A) ROC curves of the risk score for predicting 1-year, 3-year, and 5-year survival. (B) ROC curves of the risk score combined with clinical factors for predicting 1-year, 3-year, and 5-year survival. (C) Forest plot of the prognostic impact of 18 variables. (D) Scatterplots illustrate the distribution of risk score and survival status of CRC patients. (E) Kaplan-Meier curves show the independent relevance between overall survival time and risk score.

Subsequently, to evaluate the influence of clinical characteristics on overall survival, the previous model was further combined with three clinical characteristics (age, gender and pathologic stage) to construct a new Cox model (**Table S8**). As shown in **Figure 5C**, variables were significantly expect gender (*P* > 0.05). The average concordance index (C-index) of new model was 0.842 and the average AUC values predicted by the new model for 1-year, 3-year, and 5-year were 0.865, 0.839, 0.914, respectively (**Figure 5B**), the performance of this model was slightly improved compared to the previous model. We calculated the risk score of each sample based on the model that incorporates clinical characteristics and grouped samples by median risk score (**Figure 5D**). The results showed that the prognosis of the high-risk group was still significantly poorer than that of the low-risk group (**Figure 5E**). Therefore, a combination of 18 variables was determined as a prognostic prediction model, including 15 variables about genetic alterations and 3 clinical factors.

A prognostic nomogram for prediction of 1-year, 3-year, and 5-year overall survival based on 15 genetic alterations, age, gender and pathologic stage in CRC patients was established (**Figure S6A**). Calibration curves indicated that actual and nomogram-predicted overall survival matched very well (**Figure S6B**). For the lack of database contained these three types of data, 5-fold cross validation was performed to accomplish reliable predictive measurement. In the 5-fold cross validation, the TCGA samples were randomly divided into five different sets, four sets were used for training, and the remaining set was used for validation. As shown in **Table 2** and **Figure S7**, the average 1-year, 3-year, and 5-year AUC of the training sets were 0.871, 0.847 and 0.919, respectively, and the corresponding index of the validation sets equal to 0.861, 0.813 and 0.899, respectively.

**Table 2.** The AUC value and C-index of 5-fold cross validation

| Model | 1-Year AUC | 3-Year AUC | 5-Year AUC | C-index |
| --- | --- | --- | --- | --- |
| Training Set 1 | 0.863 | 0.818 | 0.895 | 0.843 |
| Test Set 1 | 0.854 | 0.884 | 0.954 | - |
| Training Set 2 | 0.923 | 0.863 | 0.937 | 0.886 |
| Test Set 2 | 0.688 | 0.650 | 0.830 | - |
| Training Set 3 | 0.862 | 0.838 | 0.903 | 0.839 |
| Test Set 3 | 0.856 | 0.860 | 0.988 | - |
| Training Set 4 | 0.854 | 0.851 | 0.935 | 0.837 |
| Test Set 4 | 0.934 | 0.861 | 0.799 | - |
| Training Set 5 | 0.852 | 0.866 | 0.923 | 0.834 |
| Test Set 5 | 0.973 | 0.812 | 0.924 | - |
| Average |  |  |  |  |
| Training Set | 0.871 | 0.847 | 0.919 | 0.848 |
| Test Set | 0.861 | 0.813 | 0.899 | - |

## DISCUSSION

Increasing studies have revealed the complex mechanisms of the occurrence and development of CRC, such as IncRNA-mediated regulation of Wnt/β-catenin signaling^29,30^. However, the current understanding of TME, especially the impact of TIME on the prognosis of patients with CRC, is not yet comprehensively understood.

In our study, firstly, using estimate algorithm, we calculated stromal, immune and estimate scores for CRC patients and explored the potential connection between scores and overall survival. In survival analysis between high and low score group, the result is significant in immune scores but not in stromal and estimate scores. To sum up, the tumor infiltration immune scores have a stronger clinical correlation than the stromal scores and estimate scores. Therefore, following analysis were based on TIME.

Subsequently, we used gene expression data to construct the TME of CRC patients, and compared the components of immune-infiltrating cells. Then, we explored the genetic and epigenetic alterations between different immune groups and the potential mechanism. The enrichment analysis results showed that apart from the representative immune-related biological processes such as leukocyte activation and immune response, some genetic alterations were also involved in transcription processes and neuronal signal regulation. Down-regulated genes may play a vital role in the transcription process of tumor cells and immune infiltrating cells. In addition, genes with differentially methylated sites may regulate immune responses through neural signaling pathways such as dendrite development and synapse organization, thereby affecting tumor progression and metastasis. A number of studies also reported similar findings^31-33^. Finally, a prognostic prediction model was constructed based on the selected genetic, epigenetic, demographic and clinical variables, which had a superior performance with a higher AUC or C-index value in comparison to the previous prognostic models^14,15^.

As expected, the proportion of some immune-infiltrating cells had a significantly difference between the high-immunity group and the low-immunity group. For example, M1 macrophages participate in the inflammatory response and anti-tumor immune process by secreting pro-inflammatory cytokines such as IL-1, IL-12, TNF-α and many chemokine ligands^34^, which is consistent with our findings that M1 macrophages accounted for a significantly higher proportion in the high-immune group.

Interestingly, the mutation frequency of APC, TP53, TTN and KRAS were accounted for the top four positions in both immunity cohorts, indicating that the four genes are less regulated the process related to immune infiltration but mainly participated in tumorigenesis and progression^35-37^. It is worth noting that gene mutations in different immune levels may have different effects on prognosis. For example, the mutation of TTN in the high-immunity cohort resulted in a worse prognosis for patients (*P* < 0.05), however, this situation did not occur in the high-immunity cohort. Meanwhile, a research found that TTN mutation was enriched in samples with high immunostimulatory signatures and suggested that TTN mutation load represents high TMB status^38^. Therefore, TTN mutation may be a therapeutic target for CRC patients. In contrast to the pervasive co-occurrence case, there is a unique situation in two groups (TP53-PIK3CA) that was mutually exclusive mutations, which indicates them probably share the same pathway within redundant effect.

After that, we considered whether there is a correlation between DNA methylations and gene expressions in different immune levels^39^. In general, the correlation between DNA methylations and gene expressions was more inclined to negatively correlated in the low-immunity group. Meanwhile, as shown in Figure 4D, the immune level did not affect the correlation between DNA methylations and gene expressions.

Our study provides a comprehensive interpretation of TIME for CRC patients and establishes a predictive model with superior performance. However, there are still some limitations. The first drawback is that, because lacking the multi-omics data and corresponding clinical information, we only collected data from TCGA portal and did not include data from other sources. The model built in this study cannot be verified on other datasets. The second limitation is that the application of three types of profiles used in this study, including RNA-seq, WES and DNA methylation data, which are expensive to detected and not easy to implement in clinical practice. Despite the above shortcomings, there is no denying that our research provides a new clue to explore the TIME of CRC patients in many aspects. Meanwhile, the model also has clinical prognostic value and may provide new biomarkers for the treatment of CRC patients.

## Data Availability

All data used in this study are available online.

https://portal.gdc.cancer.gov/repository

## REFERENCES

1. Siegel RL, Miller KD, Jemal A. Cancer statistics, 2020. CA Cancer J Clin. 2020;70(1):7–30.

2. Locker GY, Hamilton S, Harris J, et al. ASCO 2006 update of recommendations for the use of tumor markers in gastrointestinal cancer. J Clin Oncol. 2006;24(33):5313–5327.

3. Weitz J, Koch M, Debus J, Höhler T, Galle PR, Büchler MW. Colorectal cancer. The Lancet. 2005;365(9454):153–165.

4. Nagtegaal ID, Quirke P, Schmoll H-J. Has the new TNM classification for colorectal cancer improved care? Nat Rev Clin Oncol. 2011;9(2):119–123.

5. Nitsche U, Maak M, Schuster T, et al. Prediction of prognosis is not improved by the seventh and latest edition of the TNM classification for colorectal cancer in a single-center collective. Ann Surg. 2011;254(5):793–800; discussion 800-1.

6. Kuipers EJ, Grady WM, Lieberman D, et al. Colorectal cancer. Nat Rev Dis Primers. 2015;1:15065.

7. Ganesh K, Stadler ZK, Cercek A, et al. Immunotherapy in colorectal cancer: rationale, challenges and potential. Nat Rev Gastroenterol Hepatol. 2019;16(6):361–375.

8. Zhao P, Li L, Jiang X, Li Q. Mismatch repair deficiency/microsatellite instability-high as a predictor for anti-PD-1/PD-L1 immunotherapy efficacy. J Hematol Oncol. 2019;12(1):54.

9. Binnewies M, Roberts EW, Kersten K, et al. Understanding the tumor immune microenvironment (TIME) for effective therapy. Nat Med. 2018;24(5):541–550.

10. Hanahan D, Coussens LM. Accessories to the crime: functions of cells recruited to the tumor microenvironment. Cancer Cell. 2012;21(3):309–322.

11. Quail DF, Joyce JA. Microenvironmental regulation of tumor progression and metastasis. Nat Med. 2013;19(11):1423–1437.

12. Wei C, Yang C, Wang S, et al. Crosstalk between cancer cells and tumor associated macrophages is required for mesenchymal circulating tumor cell-mediated colorectal cancer metastasis. Mol Cancer. 2019;18(1):64.

13. Yin Y, Yao S, Hu Y, et al. The Immune-microenvironment Confers Chemoresistance of Colorectal Cancer through Macrophage-Derived IL6. Clin Cancer Res. 2017;23(23):7375–7387.

14. Zhou Y, Bian S, Zhou X, et al. Single-Cell Multiomics Sequencing Reveals Prevalent Genomic Alterations in Tumor Stromal Cells of Human Colorectal Cancer. Cancer Cell. 2020;38(6):818-828.e5.

15. Wang J, Yu S, Chen G, et al. A novel prognostic signature of immune-related genes for patients with colorectal cancer. J Cell Mol Med. 2020;24(15):8491–8504.

16. Yoshihara K, Shahmoradgoli M, Martínez E, et al. Inferring tumour purity and stromal and immune cell admixture from expression data. Nat Commun. 2013;4:2612.

17. Camp RL, Dolled-Filhart M, Rimm DL. X-tile: a new bio-informatics tool for biomarker assessment and outcome-based cut-point optimization. Clin Cancer Res. 2004;10(21):7252–7259.

18. Newman AM, Liu CL, Green MR, et al. Robust enumeration of cell subsets from tissue expression profiles. Nat Methods. 2015;12(5):453–457.

19. Robinson MD, McCarthy DJ, Smyth GK. edgeR: a Bioconductor package for differential expression analysis of digital gene expression data. Bioinformatics. 2010;26(1):139–140.

20. Koboldt DC, Zhang Q, Larson DE, et al. VarScan 2: somatic mutation and copy number alteration discovery in cancer by exome sequencing. Genome Research. 2012;22(3):568–576.

21. Mayakonda A, Lin D-C, Assenov Y, Plass C, Koeffler HP. Maftools: efficient and comprehensive analysis of somatic variants in cancer. Genome Research. 2018;28(11):1747–1756.

22. Leiserson MDM, Wu H-T, Vandin F, Raphael BJ. CoMEt: a statistical approach to identify combinations of mutually exclusive alterations in cancer. Genome Biol. 2015;16:160.

23. Morris TJ, Butcher LM, Feber A, et al. ChAMP: 450k Chip Analysis Methylation Pipeline. Bioinformatics. 2014;30(3):428–430.

24. Yu G, Wang L-G, Han Y, He Q-Y. clusterProfiler: an R package for comparing biological themes among gene clusters. OMICS. 2012;16(5):284–287.

25. Friedman J, Hastie T, Tibshirani R. Regularization Paths for Generalized Linear Models via Coordinate Descent. J Stat Softw. 2010;33(1):1–22.

26. Blanche P, Dartigues J-F, Jacqmin-Gadda H. Estimating and comparing time-dependent areas under receiver operating characteristic curves for censored event times with competing risks. Stat Med. 2013;32(30):5381–5397.

27. Therneau TM, Grambsch PM. Modeling Survival Data: Extending the Cox Model / Terry M. Therneau, Patricia M. Grambsch. Springer; 2000.

28. Jung G, Hernández-Illán E, Moreira L, Balaguer F, Goel A. Epigenetics of colorectal cancer: biomarker and therapeutic potential. Nat Rev Gastroenterol Hepatol. 2020;17(2):111–130.

29. Zhang M, Weng W, Zhang Q, et al. The lncRNA NEAT1 activates Wnt/β-catenin signaling and promotes colorectal cancer progression via interacting with DDX5. J Hematol Oncol. 2018;11(1):113.

30. Han P, Li J-W, Zhang B-M, et al. The lncRNA CRNDE promotes colorectal cancer cell proliferation and chemoresistance via miR-181a-5p-mediated regulation of Wnt/β-catenin signaling. Mol Cancer. 2017;16(1):9.

31. Alekseenko IV, Chernov IP, Kostrov SV, Sverdlov ED. Are Synapse-Like Structures a Possible Way for Crosstalk of Cancer with Its Microenvironment? Cancers (Basel).zx 2020;12(4).

32. Derniame S, Vignaud J-M, Faure GC, Béné MC. Alteration of the immunological synapse in lung cancer: a microenvironmental approach. Clin Exp Immunol. 2008;154(1):48–55.

33. Liu D, Tian S, Zhang K, et al. Erratum to: Chimeric antigen receptor (CAR)-modified natural killer cell-based immunotherapy and immunological synapse formation in cancer and HIV. Protein Cell. 2018;9(10):902.

34. Mosser DM, Edwards JP. Exploring the full spectrum of macrophage activation. Nat Rev Immunol. 2008;8(12):958–969.

35. Liu Y, Zhang X, Han C, et al. TP53 loss creates therapeutic vulnerability in colorectal cancer. Nature. 2015;520(7549):697–701.

36. Wong CC, Xu J, Bian X, et al. In Colorectal Cancer Cells With Mutant KRAS, SLC25A22-Mediated Glutaminolysis Reduces DNA Demethylation to Increase WNT Signaling, Stemness, and Drug Resistance. Gastroenterology. 2020;159(6):2163-2180.e6.

37. Zhang L, Shay JW. Multiple Roles of APC and its Therapeutic Implications in Colorectal Cancer. JNCI: Journal of the National Cancer Institute. 2017;109(8).

38. Oh J-H, Jang SJ, Kim J, et al. Spontaneous mutations in the single TTN gene represent high tumor mutation burden. npj Genom Med. 2020;5(1):33.

39. Greenberg MVC, Bourc’his D. The diverse roles of DNA methylation in mammalian development and disease. Nat Rev Mol Cell Biol. 2019;20(10):590–607.

