## Supplement figure for "Constructing Tumor Immune Microenvironment and Identifying the Immune-Related Prognostic Signatures in Colorectal Cancer Using Multi-omics Data"

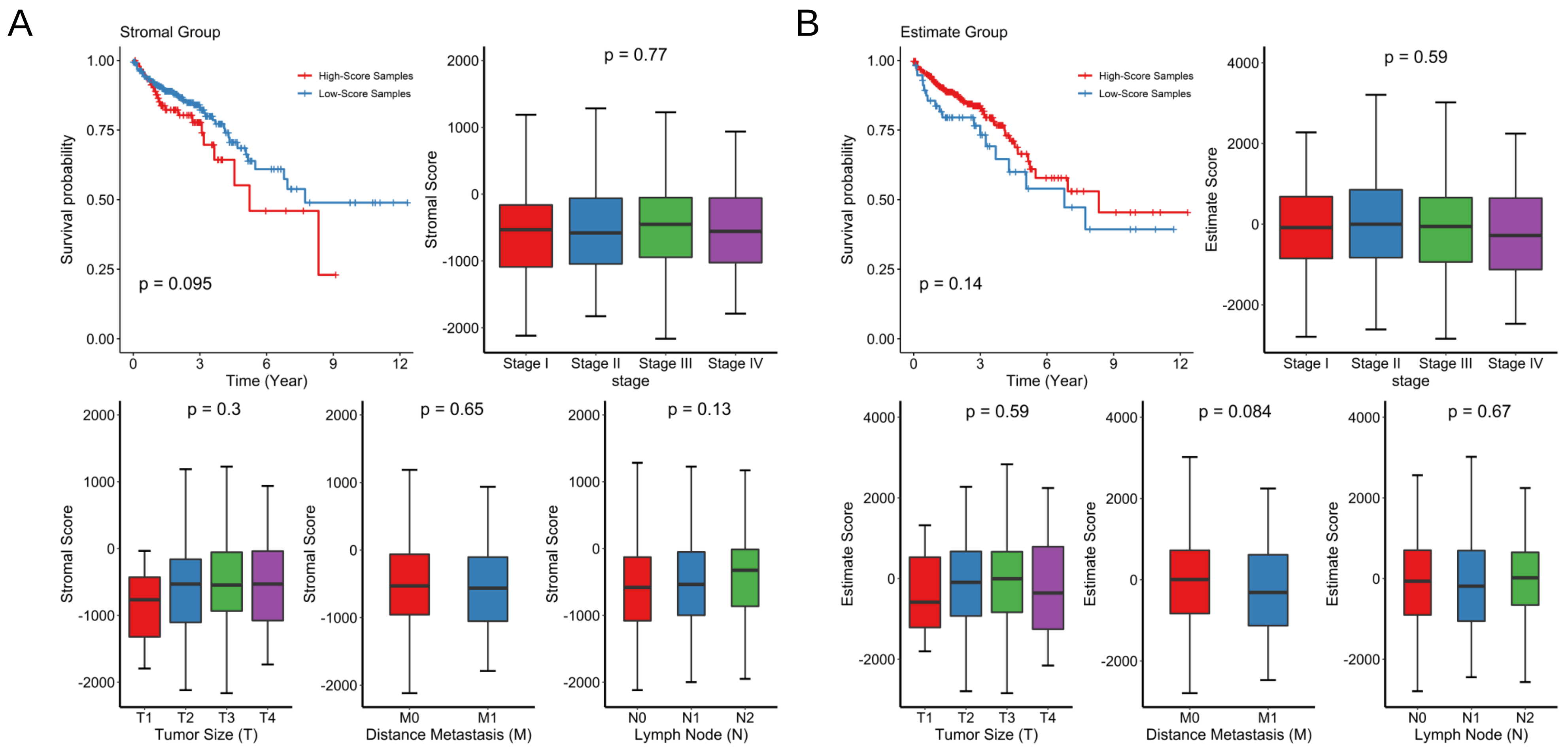


**Figure S1** The comparisons of the distributions of (A) stromal scores and (B) estimate scores on TNM stage, tumor size, distant metastasis, lymph nodes and overall survival time.

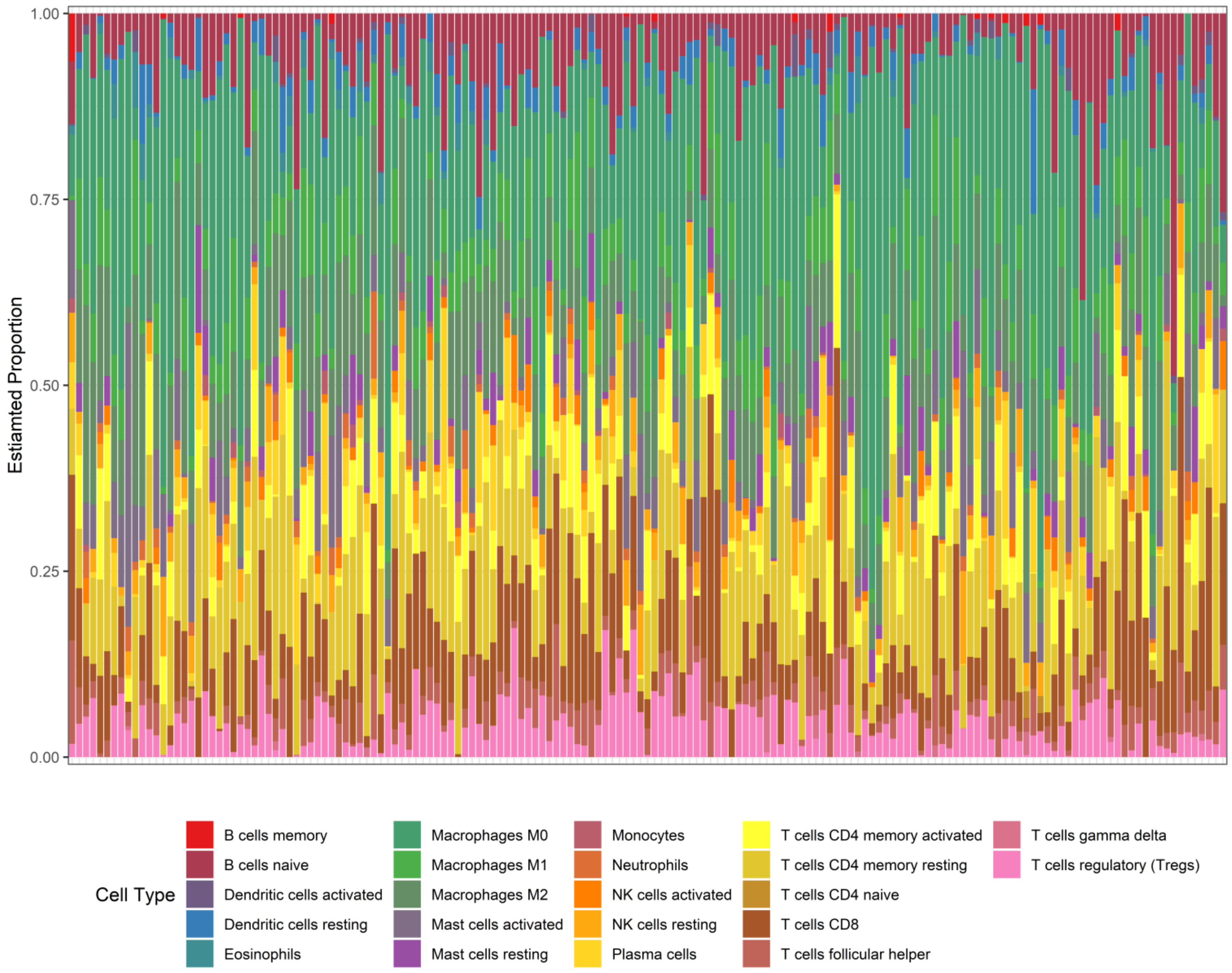


**Figure S2** The immune infiltrating cell composition of each sample with a CIBERSORT' P value less than 0.05

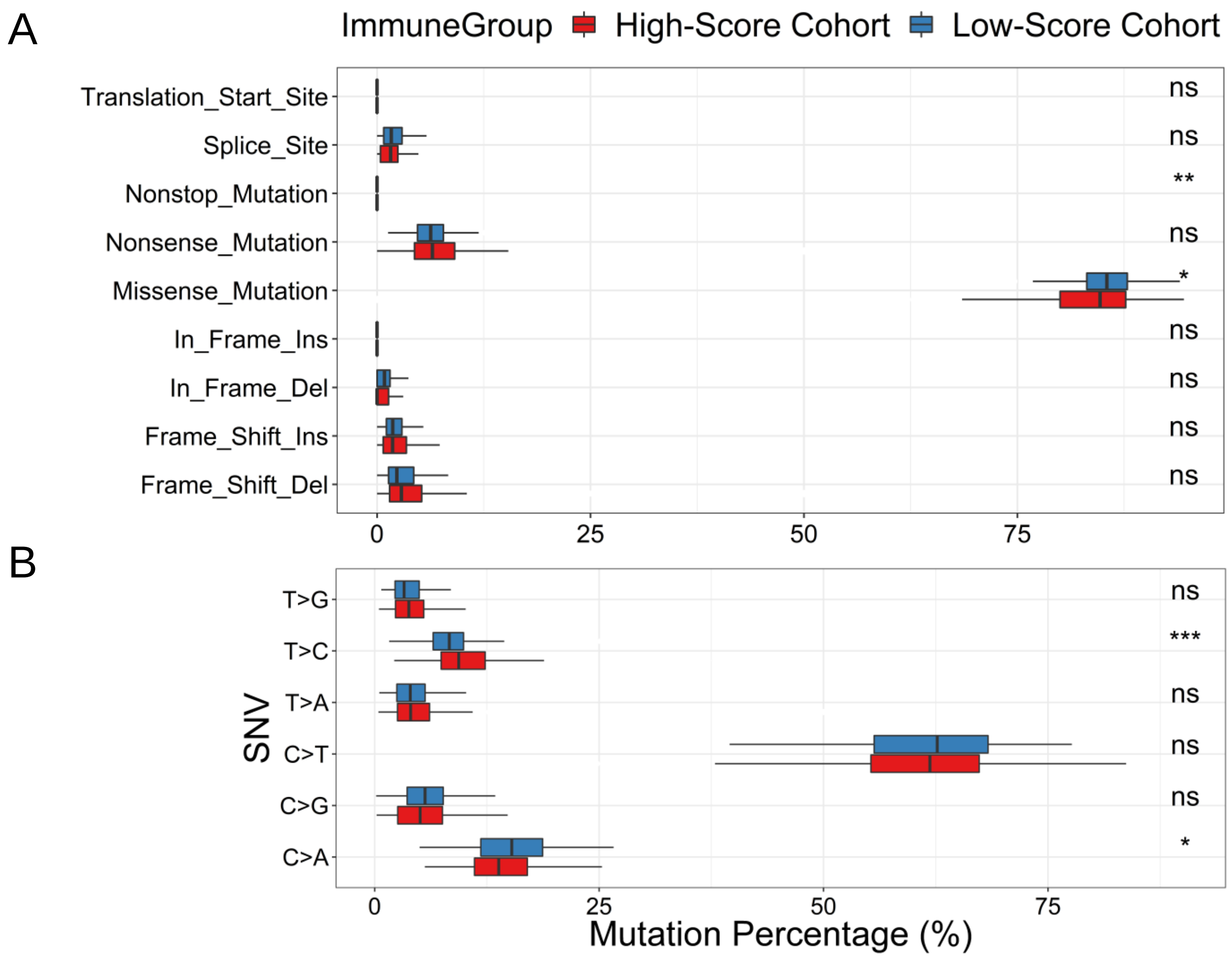


**Figure S3** The percentages of various mutation types in high-immunity and low-immunity cohorts. Boxplots respectively display the comparisons of the percentages of (A) every mutation type classified by effects, (B) SNV. Symbols indicated statistical significance for the wilcox test: ns, $P > 0.05$; *, $P \le 0.05$; **, $P \le 0.01$; ***, $P \le 0.001$.

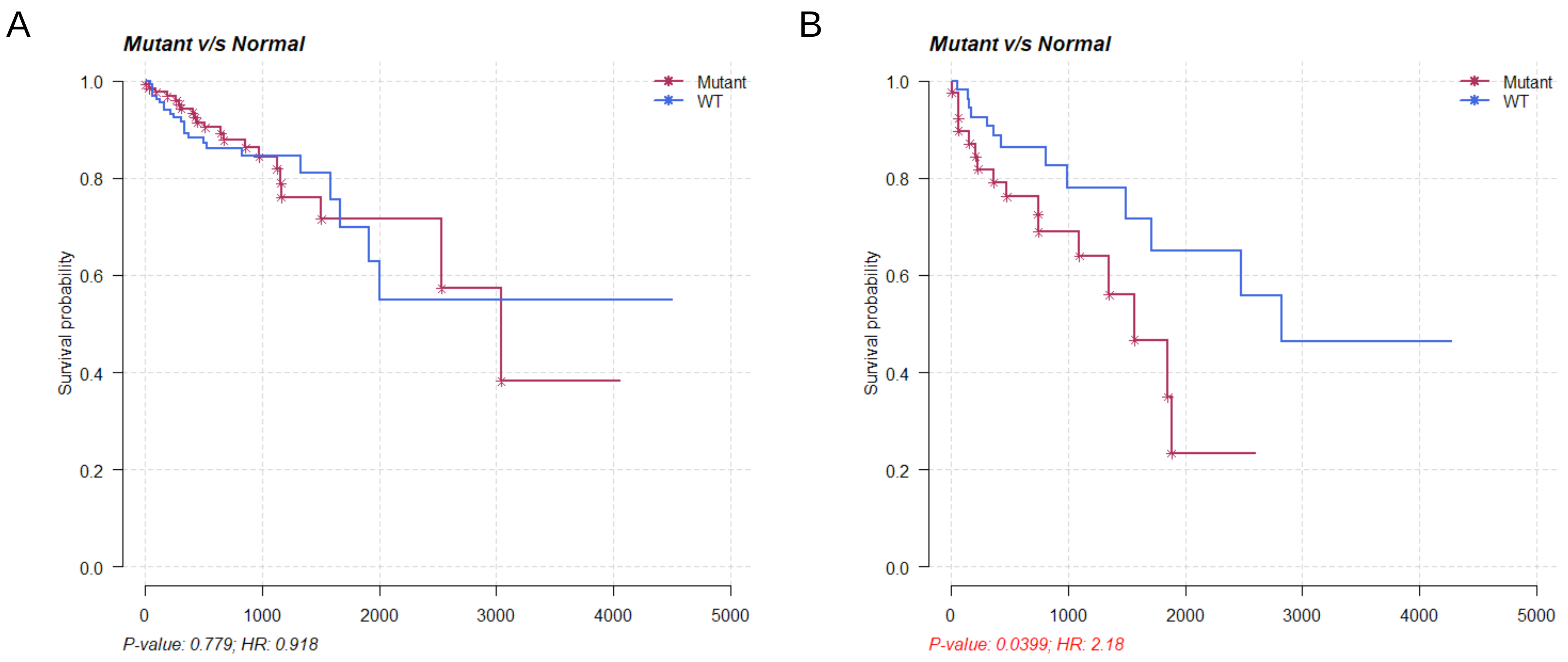


**Figure S4** Kaplan-Meier curves show the independent relevance between overall survival time and TTN mutation in (A) high-immunity group(B) low-immunity group.

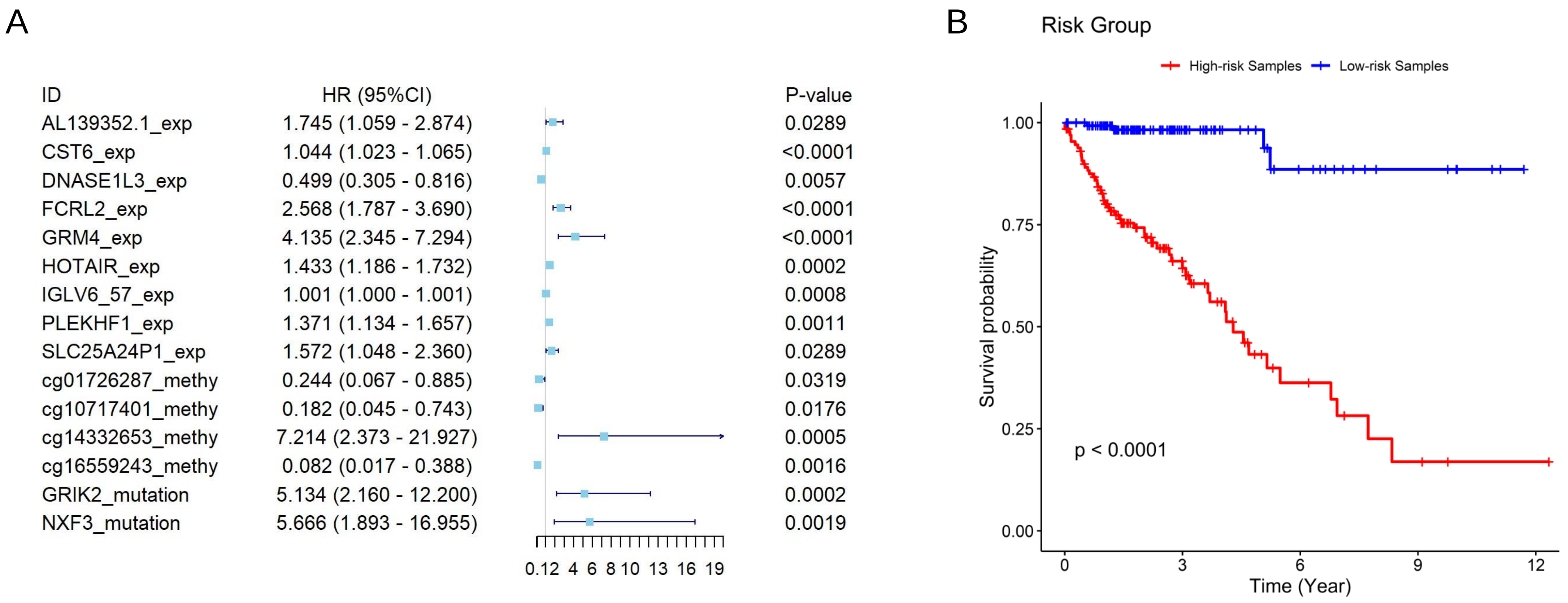


**Figure S5** (A) Forest plot of the prognostic impact of genetic and epigenetic variables. (B) Kaplan-Meier curves show the independent relevance between overall survival time and risk scores.

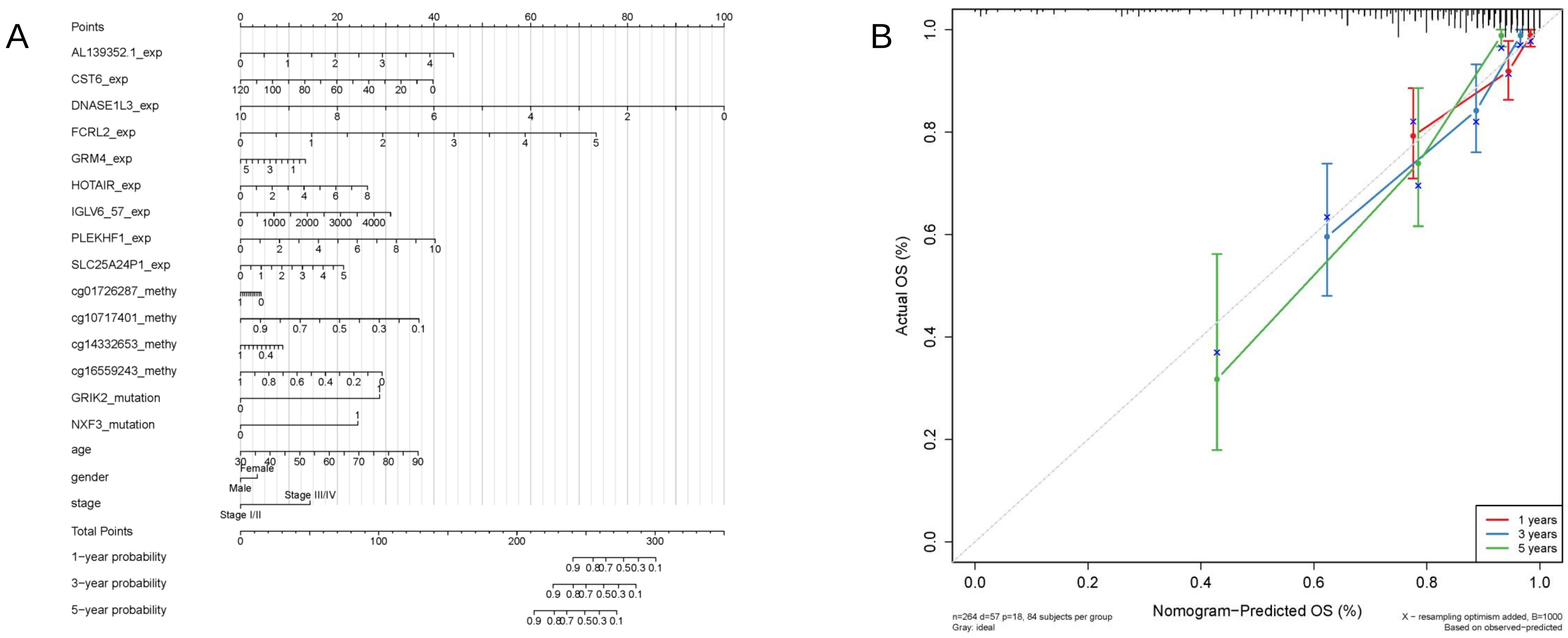


**Figure S6** Nomogram (A) and calibration plot (B) for prediction of overall survival time based on the combination of multi-omics characteristics and clinical factors in CRC.

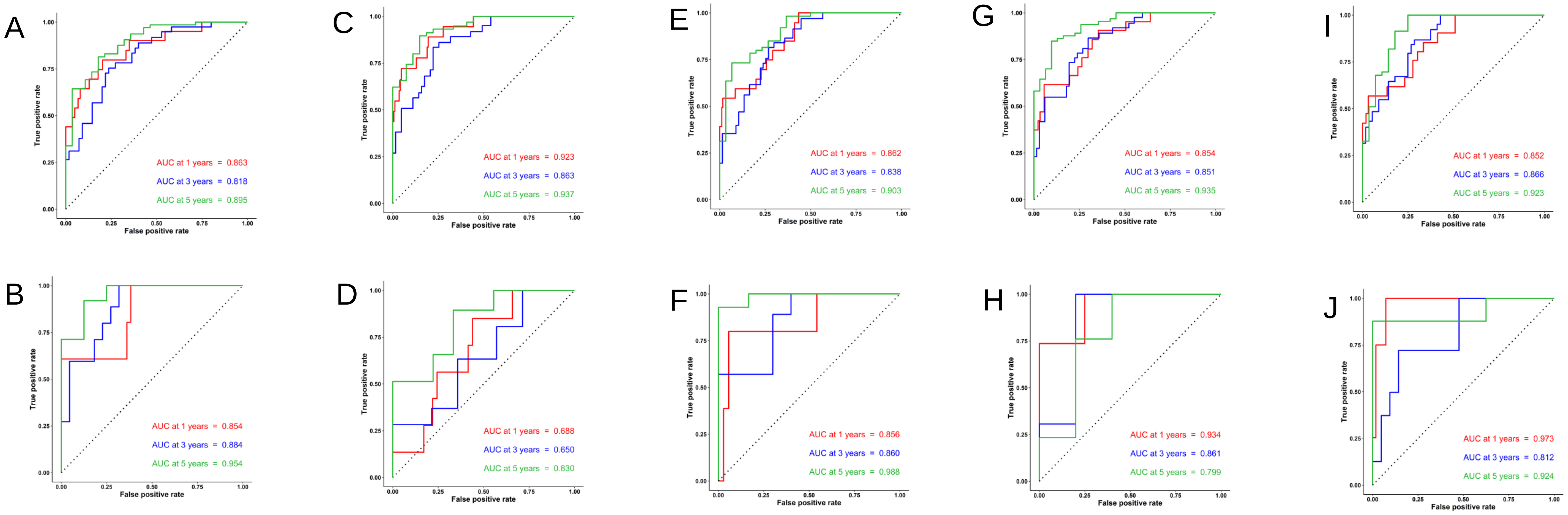


**Figure S7** The performance of the 5-fold cross validation set 1 (A-B), set 2 (C-D), set 3 (E-F), set 4 (G-H) and set 5 (I-J). Subplot (A/C/E/G/I) and (B/D/F/H/J) show the roc curves of the risk score for 1-year, 3-year and 5-year survival prediction on the training set and test set, respectively.
